# Adherence to preventive behaviors related to Covid-19 in Taiwan in 2020-2021: a population-based follow-up survey

**DOI:** 10.1101/2025.03.04.25323269

**Authors:** Chieh-Yin Wu, Chih-Chan Lan, Shu-Wan Jian, Chih-Chi Yang, Wen-Chi Hsu, Pau-Chung Chen, Shu-Sen Chang, Hsien-Ho Lin, the Taiwan COVID-19 Prevention Behaviors Survey Team

## Abstract

**Objectives:** The COVID-19 pandemic was associated with significant changes in preventive behaviors worldwide. We conducted a cohort follow-up survey to understand these behavioral changes in Taiwan during 2020 and 2021.

**Methods:** A population-based survey with three follow-up rounds was implemented among individuals aged 20 and older. The survey included questions about perceptions of COVID-19 risk and adherence to preventive behaviors promoted by the Taiwan CDC in its New Life Strategies.

**Results:** Between July and September 2020, a period with no local COVID-19 cases, adherence to indoor and outdoor social distancing was 51.5% and 63.5%, respectively, while 81.4% of people indicated they would wear facial masks if social distancing could not be practiced. In stratified analysis, females and the elderly exhibited significantly higher adherence to most of the preventive measures. We observed an increase in adherence to preventive behaviors from 2020 to the middle of 2021, with no signs of fatigue.

**Conclusions:** The survey provided critical empirical evidence on the adherence to preventive behaviors in the general population in Taiwan during the first two years of the pandemic. As non-pharmaceutical interventions (NPI) are essential measures during the early stage of any pandemic, we believe that regular monitoring of preventive behaviors could serve as a foundation for pandemic preparedness. More efforts are needed to identify the most feasible and cost-effective methods for surveillance of preventive behaviors.

## Background

The COVID-19 pandemic first emerged in late 2019 and swiftly spread across the globe in early 2020, being declared a pandemic by March of that year. In Taiwan, the situation remained relatively stable initially, with only sporadic cases linked to imported cases, or, at times, no local cases reported throughout the entirety of 2020 and early 2021. It wasn’t until April 20, 2021, that Taiwan recorded its first local case after an uninterrupted of 121-day streak without any local transmissions. A local epidemic emerged from this point, proven to be more significant in scale than earlier occurrences and persisted for approximately two months (as depicted in Figure 1).

**Figure 1.**
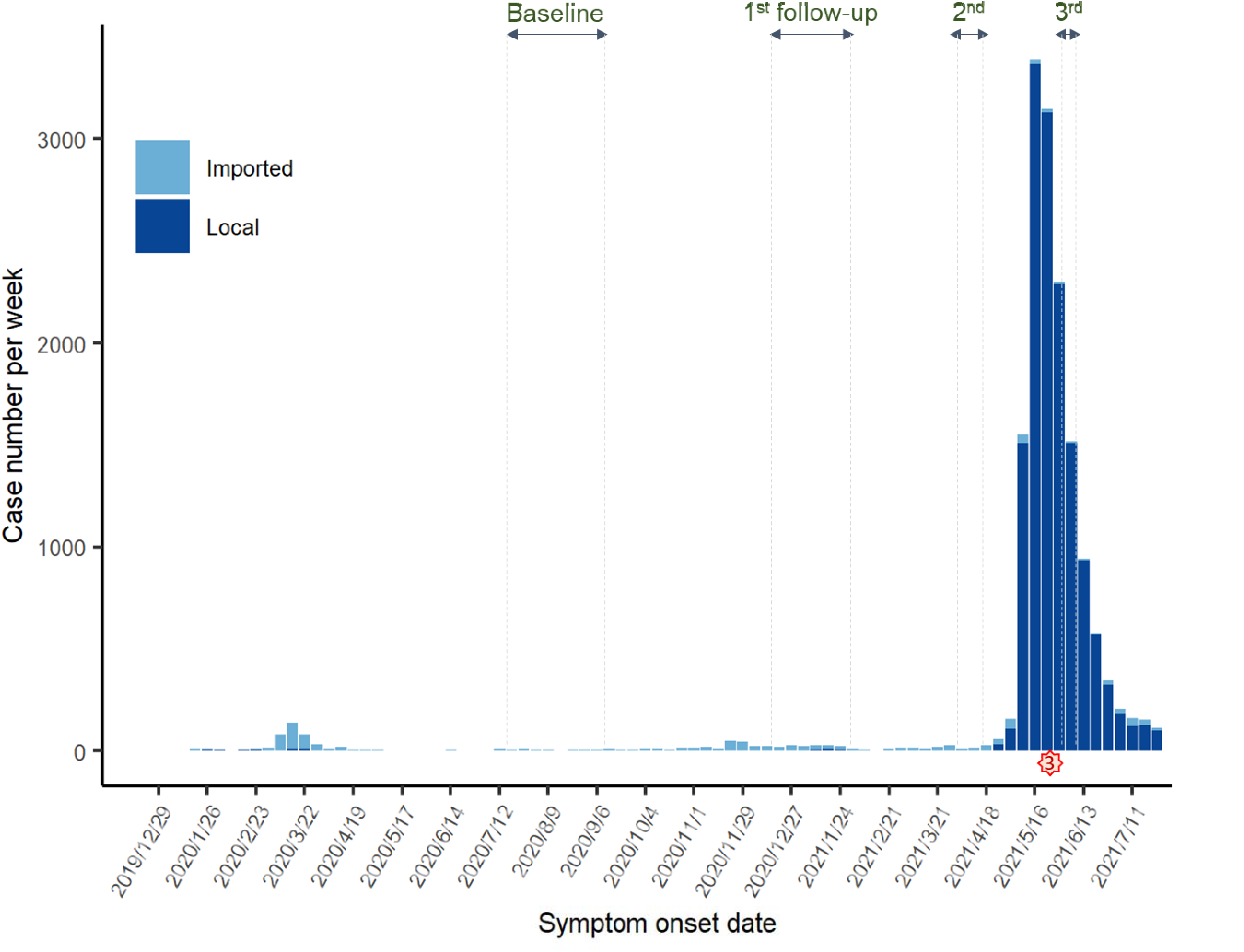
Epidemic trend of COVID-19 in Taiwan and the corresponding time periods when the baseline survey and three waves of follow-up surveys were conducted.

Subsequently, a nationwide level 3 alert was declared, and various enhanced restrictions were implemented, including the suspension of public venues, from May 19 to July 26, 2021.

The Taiwan government introduced the “New Life Strategies for Epidemic Prevention” [1] on April 30, 2020. This strategy aimed to reduce both the contact rate and infection rate, promoting measures such as extended social distancing, increased handwashing, and mask-wearing under specific conditions. These actions were crucial in preventing virus transmission and flattening the peak of the epidemic. They were collectively referred to as critical non-pharmaceutical interventions (NPIs).

Recognizing the importance of monitoring these preventive behaviors, knowledge, and public perceptions of risk, the WHO Regional Office for Europe launched a standard protocol [2] in March 2020, providing guidance on implementing such monitoring systems. Since then, many European countries initiated their own COVID-19 Snapshot Monitoring (COSMO) and rapidly conducted several surveys based on the protocols. The UK’s Department of Health and Social Care also conducted bi-weekly or monthly surveys [3] related to COVID-19 symptoms and preventive behaviors. Beyond Europe, countries like Ethiopia and Japan assessed preventive behavior through surveys. The World Bank initiated monthly phone-based surveys to monitor COVID-19’s effects on firms and households in Ethiopia [4] since April 2020. The Japanese government collaborated with LINE Corporation to distribute several rounds of national-scale surveys [5, 6].

In Taiwan, some researchers investigated the adoption of preventive behavior during the pandemic. However, most of these studies focused on specific populations such as the elderly, healthcare workers, and people with mental illnesses [7-9]. While these studies aimed to identify factors associated with preventive behaviors, their participants might not have been entirely representative of the general population.

Moreover, the survey samples, collected cross-sectionally through social media like Facebook advertisements or telephone interviews [10-12], might not capture behavioral changes over time due to the evolving nature of the pandemic. In contrast to the comprehensive monitoring systems in Europe, Taiwan lacked a routine system to consistently measure and examine the public’s implementation of preventive behaviors. Recognizing this gap, under the delegation of Taiwan Centers for Disease Control (CDC), we conducted multiple rounds of surveys to gain insights into the compliance with key preventive behaviors and to track the trends in behavioral changes over time among the general population in Taiwan.

## Method

### Study Protocol

Participants were recruited from community-based settings using a multi-stage, stratified, cluster sampling design at the city level among individuals aged 20 years and older. The anticipated sample sizes in each city/county were calculated based on the city/county’s population and the distribution of gender and age groups (20-34, 35-49, 50-64, and 65+ years). This followed the principle of probability proportional to size. In cases where certain strata were allocated a sample size of fewer than 100, we intentionally oversampled to ensure a minimum sample size of 100, enabling a more in-depth analysis.

Trained interviewers conducted the baseline survey using web-based questionnaire on tablet computers in face-to-face interviews from July to September, 2020. All interviewers were registered on our online platform, constructed on Microsoft Teams and monitored by project research assistants, to streamline the process. The questionnaire was created using Microsoft Forms and incorporated into the online platform. Every submitted response was instantly updated to a secure database accessible to the members of the core research team.

Potential participants were approached at various public locations frequently visited by the residents, including local traditional markets, temples, churches, gyms, public libraries, public health centers, and university campuses. After obtaining the consent, the interviewers conducted interviews lasting between 20 to 25 minutes. Each interview location was overseen by at least one supervisor responsible for monitoring the interview process. These supervisors also conducted random sample reinspections by calling the participants. Participants who gave consent to be re-contacted were invited to participate in follow-up surveys (details provided in the Appendix). Three waves of follow-up surveys were conducted, either through telephone interviews or web-based self-administered questionnaires, over the following periods: (1)

December 14, 2020, to February 8, 2021, (2) March 27 to April 19, 2021, (3) June 7 to June 14, 2021. The questionnaire used in these surveys were designed to assess preventive behaviors, health beliefs, attitude towards the COVID-19, and participants’ health conditions. The section on preventive behaviors covered those outlined in the New Life Strategies announced by the CDC, including hand washing, disinfection measures, social distancing, etc. Response options were configured to capture the frequency of each behavior over the past seven days, utilizing a 5-point Likert Scale that ranged from “always” to “never”. The questionnaire used in the follow-up surveys was a condensed version of the one used in the baseline survey.

### Data Analysis

To ensure our results are nationally representative, we applied weighting to the data based on gender and age distributions within each city/county. These distributions, reflective of the population as of May 2020, were extracted from the website of the Department of Household Registration, Ministry of the Interior. The weights were calculated as follows –

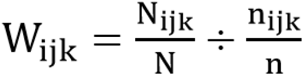

*Wijk*: The weight of people in city i, gender j and age group k.

*Nijk*: In city i and gender j, the number of total population in age group k

*nijk*: In city i and gender j, the number of completed samples in age group k

*n*: Number of completed and included samples

*N*: Number of population nationally

We calculated the weighted percentage of participants executing each behavior across the 5-level frequency scale. To assess changes over time, we compared results from baseline to the third follow-up using the Friedman test and Wilcoxon Signed Rank test.

### Ethical Statement

This study was approved by the Research Ethics Committee of National Taiwan University (202007HM008).

## Results

### Participants

Out of the initial 4,954 participants enrolled at baseline, a total of 4,788 individuals (96.7%) were included in the analysis after excluding those who had missing residential area data or had data not meeting the quality check criteria (Figure 2). Among the included participants, 44.4% were male, 44.6% were aged above 50 years, and 48.3% held a college degree. When considering the distribution of included participants across various cities/counties, it closely mirrored the demographic composition of the general population in Taiwan in terms of sex and age groups (refer to Appendix Table 2). Throughout each wave of follow-up surveys, we observed a higher proportion of female participants, those of a younger age, and those with a higher education level (Appendix Table 1).

**Figure 2.**
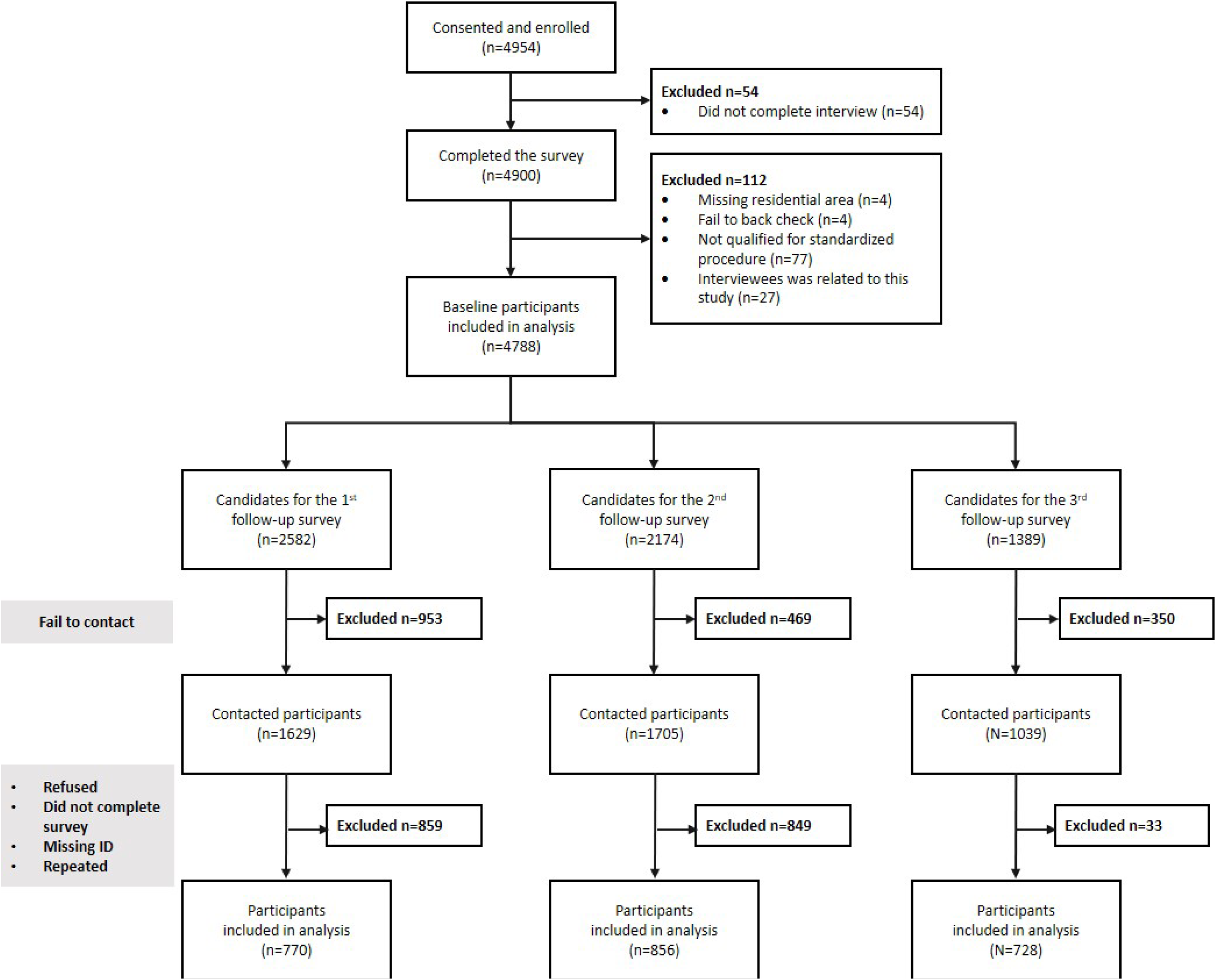
Flowchart of participants in the baseline the follow-up surveys.

### Preventive behaviors

We summarized the practices of each preventive behavior by calculating the percentage of weighted results from the baseline cohort, focusing on individuals who reported preforming these behaviors ‘often’ or ‘always’. The percentage of people who practiced washing hands with water and soap was 77.4% (Table 2). Those who covered their mouth and face when coughing or sneezing and avoided touching their eyes and nose without having washed their hands were 77.2% and 63.5%, respectively. Social distancing was practiced by 51.5% of individuals indoors and 63.5% outdoors, with 81.4% indicating a willingness to wear masks when social distancing was not feasible. Additionally, 42.0% and 54.4% of people avoided using public transportation and visiting crowded places, respectively. During the period between July and September 2020, 32.2% of respondents cancelled their plans for international travel due to the pandemic. The majority of participants agreed that the epidemic was well-controlled in Taiwan, with only 34.77 % and 17.88% of them were worried about the insufficiency of the medical resources and living supplies, respectively. 83.4% of the population had confidence in the government’s outbreak control strategies. While people generally had faith in their own adherence to preventive behaviors, 11.6% expressed varying degrees of doubt or lack of confidence in the practices of other people.

**Table 1.**
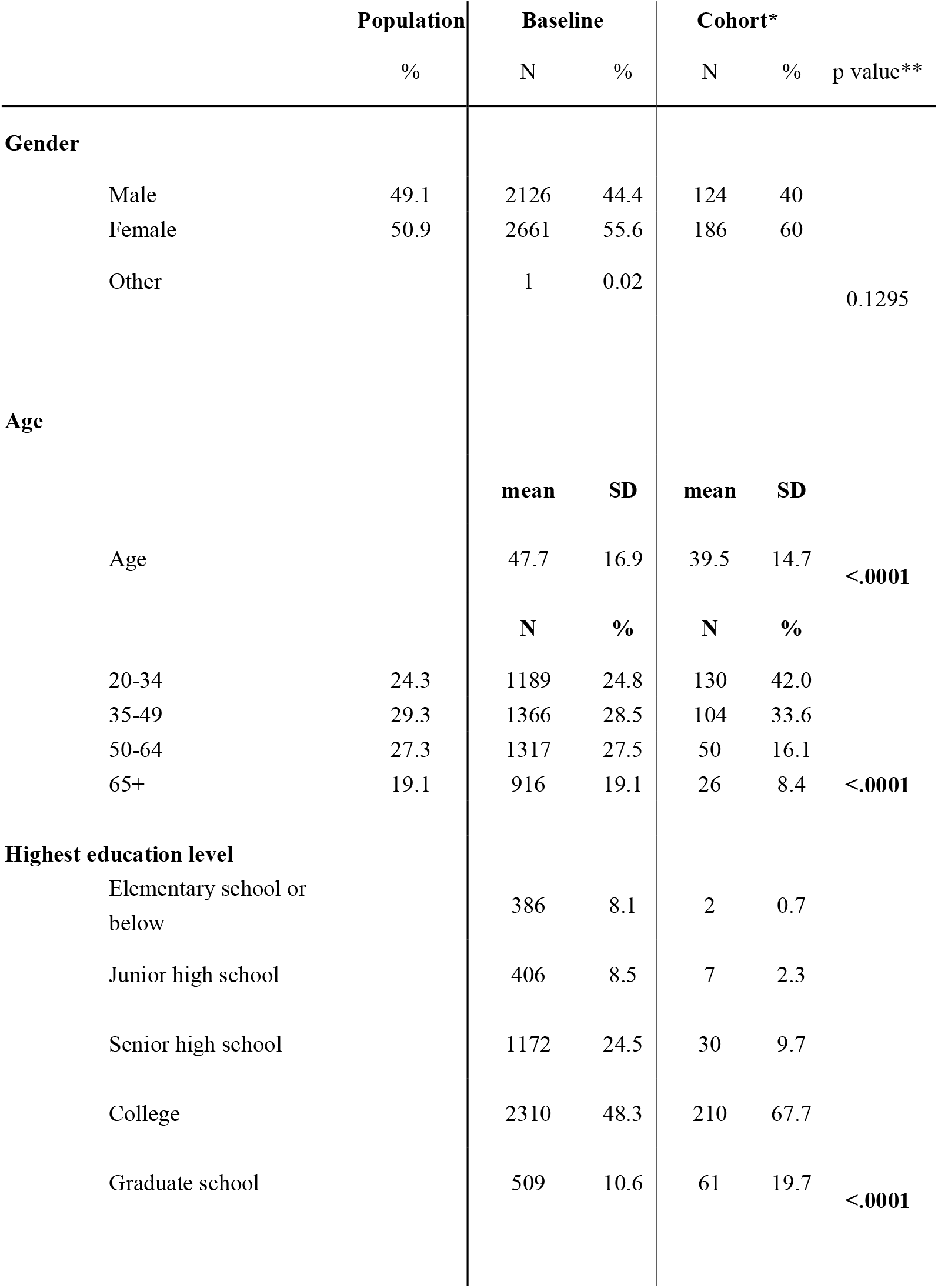
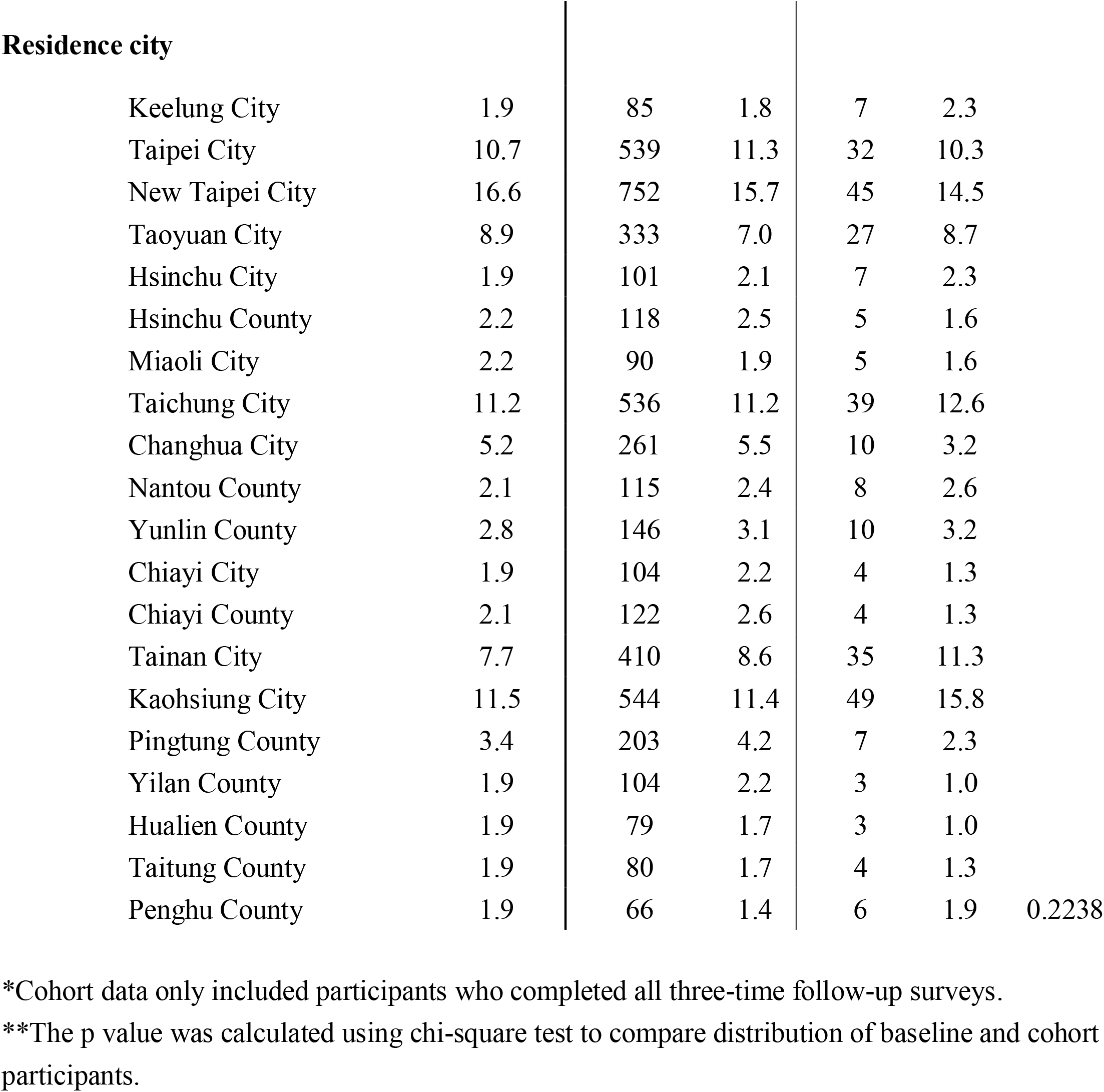
Demographic distribution of Taiwan population, participants of baseline and cohort survey.

|  |  | Population | Baseline |  | Cohort* |  | p value** |
| --- | --- | --- | --- | --- | --- | --- | --- |
|  |  | % | N | % | N | % |  |
| **Gender** |  |  |  |  |  |  |  |
|  | Male | 49.1 | 2126 | 44.4 | 124 | 40 |  |
|  | Female | 50.9 | 2661 | 55.6 | 186 | 60 |  |
|  | Other |  | 1 | 0.02 |  |  | 0.1295 |
| **Age** |  |  |  |  |  |  |  |
|  |  |  | **mean** | **SD** | **mean** | **SD** |  |
|  | Age |  | 47.7 | 16.9 | 39.5 | 14.7 | <.0001 |
|  |  |  | **N** | **%** | **N** | **%** |  |
|  | 20-34 | 24.3 | 1189 | 24.8 | 130 | 42.0 |  |
|  | 35-49 | 29.3 | 1366 | 28.5 | 104 | 33.6 |  |
|  | 50-64 | 27.3 | 1317 | 27.5 | 50 | 16.1 |  |
|  | 65+ | 19.1 | 916 | 19.1 | 26 | 8.4 | <.0001 |
| **Highest education level** |  |  |  |  |  |  |  |
|  | Elementary school or below |  | 386 | 8.1 | 2 | 0.7 |  |
|  | Junior high school |  | 406 | 8.5 | 7 | 2.3 |  |
|  | Senior high school |  | 1172 | 24.5 | 30 | 9.7 |  |
|  | College |  | 2310 | 48.3 | 210 | 67.7 |  |
|  | Graduate school |  | 509 | 10.6 | 61 | 19.7 | <.0001 |

| Residence city |  |  |  |  |  |  |
| --- | --- | --- | --- | --- | --- | --- |
| Keelung City | 1.9 | 85 | 1.8 | 7 | 2.3 |  |
| Taipei City | 10.7 | 539 | 11.3 | 32 | 10.3 |  |
| New Taipei City | 16.6 | 752 | 15.7 | 45 | 14.5 |  |
| Taoyuan City | 8.9 | 333 | 7.0 | 27 | 8.7 |  |
| Hsinchu City | 1.9 | 101 | 2.1 | 7 | 2.3 |  |
| Hsinchu County | 2.2 | 118 | 2.5 | 5 | 1.6 |  |
| Miaoli City | 2.2 | 90 | 1.9 | 5 | 1.6 |  |
| Taichung City | 11.2 | 536 | 11.2 | 39 | 12.6 |  |
| Changhua City | 5.2 | 261 | 5.5 | 10 | 3.2 |  |
| Nantou County | 2.1 | 115 | 2.4 | 8 | 2.6 |  |
| Yunlin County | 2.8 | 146 | 3.1 | 10 | 3.2 |  |
| Chiayi City | 1.9 | 104 | 2.2 | 4 | 1.3 |  |
| Chiayi County | 2.1 | 122 | 2.6 | 4 | 1.3 |  |
| Tainan City | 7.7 | 410 | 8.6 | 35 | 11.3 |  |
| Kaohsiung City | 11.5 | 544 | 11.4 | 49 | 15.8 |  |
| Pingtung County | 3.4 | 203 | 4.2 | 7 | 2.3 |  |
| Yilan County | 1.9 | 104 | 2.2 | 3 | 1.0 |  |
| Hualien County | 1.9 | 79 | 1.7 | 3 | 1.0 |  |
| Taitung County | 1.9 | 80 | 1.7 | 4 | 1.3 |  |
| Penghu County | 1.9 | 66 | 1.4 | 6 | 1.9 | 0.2238 |

\*Cohort data only included participants who completed all three-time follow-up surveys.
\*\*The p value was calculated using chi-square test to compare distribution of baseline and cohort participants.

**Table 2.** The prevalence of COVID-19 practicing preventive behaviors in baseline survey, overall and in social demographic groups.

|  | Wash hands with water and soap |  | Wash hands with alcohol sanitizer |  | Avoid touching eyes and nose |  | Cover mouth when sneezing or coughing* |  | Keep indoor social distancing |  | Keep outdoor social distancing |  | Wear mask when being unable to keep social distancing |  |
| --- | --- | --- | --- | --- | --- | --- | --- | --- | --- | --- | --- | --- | --- | --- |
|  | % | p value | % | p value | % | p value | % | p value | % | p value | % | p value | % | p value |
| **Overall** | 77.38 |  | 59.29 |  | 63.49 |  | 77.2 |  | 51.46 |  | 63.48 |  | 81.47 |  |
| **Gender** |  |  |  |  |  |  |  |  |  |  |  |  |  |  |
| Male | 71.91 |  | 53.2 |  | 60 |  | 73.8 |  | 48.69 |  | 61.26 |  | 75.61 |  |
| Female | 82.69 | <.0001 | 65.02 | <.0001 | 66.88 | <.0001 | 80.61 | <.0001 | 54.15 | 0.0002 | 65.64 | 0.0017 | 87.15 | <.0001 |
| **Age group** |  |  |  |  |  |  |  |  |  |  |  |  |  |  |
| 20-34 | 71.79 |  | 56.16 |  | 60.64 |  | 82.18 |  | 36.43 |  | 51.82 |  | 76.62 |  |
| 35-49 | 78.99 |  | 62.22 |  | 67.8 |  | 81.47 |  | 51.47 |  | 62.51 |  | 82.01 |  |
| 50-64 | 78.66 |  | 58.6 |  | 63.44 |  | 74.12 |  | 55.59 |  | 67.21 |  | 82.65 |  |
| 65+ | 80.24 | <.0001 | 59.93 | 0.0292 | 60.48 | <.0001 | 64.89 | <.0001 | 65.07 | <.0001 | 74.85 | <.0001 | 85.19 | <.0001 |
| **Highest education level** |  |  |  |  |  |  |  |  |  |  |  |  |  |  |
| Elementary school and below | 76.28 |  | 57.52 |  | 51.41 |  | 54.26 |  | 59.53 |  | 69.69 |  | 83.07 |  |
| Junior high school | 74.55 |  | 53.29 |  | 51.23 |  | 66.02 |  | 61.24 |  | 65.54 |  | 82.07 |  |
| Senior high school | 77.52 |  | 59.28 |  | 62.03 |  | 72.38 |  | 56.35 |  | 66.38 |  | 81.75 |  |
| College | 78.17 |  | 60.85 |  | 66.81 |  | 82.47 |  | 47.12 |  | 60.79 |  | 81.14 |  |
| Graduate school | 76.52 | 0.5311 | 57.65 | 0.1083 | 70.17 | <.0001 | 84.79 | <.0001 | 46.57 | <.0001 | 62.99 | 0.001 | 80.63 | 0.882 |

(\*34.8% of participants who replied not applicable were removed)

Females exhibited higher adherence to all of the major preventive behaviors than males. When comparing different age groups, the youngest group (aged 20-34 years) exhibited the lowest adherence to these measures, with the exception of covering the mouth when sneezing and coughing. In contrast, the oldest group (aged 65+) demonstrated the highest adherence rates for behaviors such as washing hands, maintaining social distance, and wearing masks (Table 2).

### Behavioral change

To assess behavioral changes over time, we analyzed data for a subset of 310 participants who responded to all three waves of follow-up surveys. There was evidence for a change in the prevalence of most preventive behaviors over the study period (p-value < 0.001). Initially, there was an increase in the prevalence of most preventive behaviors between the baseline and the first wave of follow-up survey (Figure 3). In the second and third waves of follow-up survey, there was a decline in these practices and a marked increase to the highest prevalence, respectively. For hand washing, the percentage of individuals using soup decreased slightly from 77.9% to 74.8% (p-value = 0.023), while the percentage of those using hand sanitizer increased from 51.9% to 64.5% (p-value < 0.0001), in the second and third waves, respectively. The practice of covering mouth and nose while sneezing showed a substantial increase from 54.5% to 86.2% (p-value < 0.0001). As for maintaining social distancing, the practice of indoor social distancing slightly raised but declined in the last follow-up survey, while the practice of outdoor social distancing followed a similar trend. Nevertheless, the use of face masks when physical distancing was not feasible remained high, increasing from 84.8% to 99.7%.

**Figure 3.**
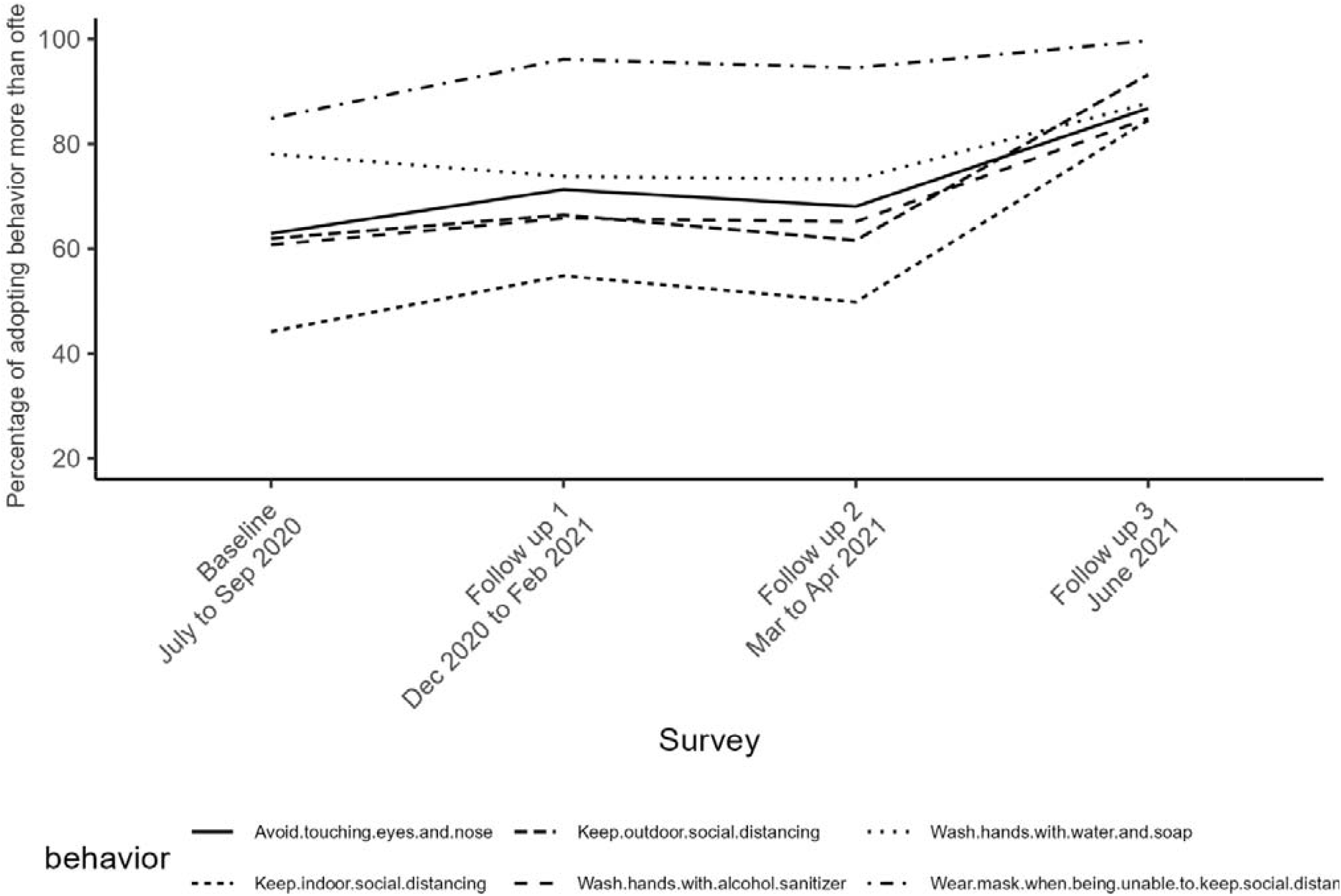
Trends in the prevalence of practicing six protective behaviors across survey waves.

## Discussion

Our study marks the first large-scale population-based survey in Taiwan assessing adherence to preventive behaviors during the COVID-19 pandemic. The baseline survey offered insights into behavior practices in the general population at a time when nearly no local epidemic transmission occurred. Across three follow-up surveys, we observed significant increases in adherence to preventive behaviors, although with some variations under different disease burden conditions. Generally, most participants reported performing preventive behaviors “more than often”, with over 60% adherence.

One major factor potentially affecting the adoption of preventive behaviors was the perception of risk, closely tied to the local epidemic level. Figure 1 illustrates the timing of each survey relative to the local and global COVID-19 burden. Initially, Taiwan had effectively controlled the epidemic, with no local and very few imported cases. The first follow-up survey occurred during a global epidemic peak, coinciding with a slight increase in Taiwan’s imported cases. The second and third follow-up surveys were implemented before and after the severe local outbreaks from May to July 2021. Compared to the baseline, we noticed that the percentage of people adopting preventive behaviors “more than often” increased with the international epidemic, decreased during a relatively quiet disease burden period, but eventually climbed to a comparably high level during the local epidemic peak. The increased preventive behavior observed from the baseline to the first and second follow-up rounds could have contributed to the effective control during the global alpha strain epidemic. However, it also implies that the level of NPI implemented at that time might not have been robust enough to prevent outbreaks between the second and third follow-up rounds.

Additionally, we compared our results with two other online surveys delivered through LINE (see Appendix). These cross-sectional online surveys, though limited in the number of questions due to LINE platform’s restrictions, quickly garnered a large number of responses. They provided additional estimates and trends regarding adherence to major preventive behaviors. Almost all of the preventive behaviors either remained stable or increased over time from Aug 2020 to late Jan 2021(see Appendix Figure). This trend aligns with the results from our follow-up cohort survey, providing further confirming the robustness of our findings.

Numerous surveys have been conducted in Taiwan since the onset of the COVID-19 epidemic. However, to our knowledge, none have investigated the adherence to preventive behavior in the general population. Most studies have either discussed the association between preventive behaviors and various factors [13, 14] or primarily focused on specific populations, e.g., health care workers[15]. Globally, direct comparisons of compliance between countries have been challenging due to differences in survey questions and scenarios. Nevertheless, studies in Cameroon, France, USA and some Asian countries indicated higher adherence to preventive behaviors among females and older individuals [16-19], consistent with our observations. In Singapore, married individuals were found to be more likely to adopt preventive behaviors [20]. It is worth noting that most of these surveys were conducted online through specific platforms (e.g., Facebook or media) and employed a cross-sectional design, contrasting with our study’s approach.

Our baseline survey utilized clustered sampling instead of a fully random sample, potentially introducing sampling bias. In addition, we specifically recruited some participants form locations like schools and health-related facilities, which could lead to selection bias. It is noteworthy that the proportion of participants with an education level above college in our baseline survey (59.3%) was slightly higher than that in the 2017 Taiwan Social Change Survey (48.7%). There’s a possibility that those who consented to participate in follow-up surveys had heightened health awareness and were more committed to executing preventive behaviors. Furthermore, participants who underwent in-person interviews might have been predisposed to provide socially desirable responses, potentially introducing social desirability bias into our findings. Due to these factors, our study might overestimate the compliance with preventive behaviors in Taiwan. However, we emphasized avoiding encouragement or leading questions in the written protocols and interviewer training to mitigate favorable responses and to ensure the accuracy and reliability of our data.

To our knowledge, this study was the only survey conducted in person with a nationally representative design. This collaborative effort involved the Taiwan CDC, the Taiwan Public Health Association and eleven public health schools across Taiwan. The extensive network and dedicated team enabled us to swiftly execute face-to-face interviews on a national scale. Through the interviews, we were able to administer a comprehensive questionnaire that not only address the primary research objectives but also delved into various critical factors that could influence behavior. The in-person interview approach played a pivotal role in achieving a high response rate during the follow-up rounds of our study. The cohort design, which involved tracking the same group of participants over time, stands as one of the most significant strengths. This approach allowed us to capture behavioral changes within the same population. While it is important to acknowledge that our findings may primarily represent certain demographic groups, which were younger and with higher levels of education in our follow-up samples, it’s worth noting that the observed trends within this population were not influenced by other factors. In contrast, more conventional survey designs, such as multiple cross-sectional surveys, can estimate the prevalence of adherence at a national or regional level during each round. However, when it comes to analyzing the trend in behavior, these estimates may be affected by the distribution of the sampled population. Furthermore, our team is working on additional analyses to understand the pattern and characteristics of behavioral changes. These analyses, which are unique to follow-up cohort design, will provide valuable insights into the dynamics of preventive behavior changes over time.

The results of this study provided a comprehensive understanding of the adherence to preventive behaviors among the general population in Taiwan. Moreover, they shed light on potential behavioral changes across various epidemic levels. Given the importance of non-pharmaceutical interventions (NPIs) in managing the ongoing COVID-19 epidemic and potential future pandemics, we believe that regular monitoring of preventive behaviors can yield critical insights and aid in pandemic control efforts. Therefore, more efforts are warranted to identify the most feasible and cost-effective methods for surveillance of preventive behaviors.

## Data Availability

All data produced in the present study are available upon reasonable request to the authors

## Acknowledgement

We thank Taiwan Public Health Association for assisting this project and training sessions for interviewers. We also thank the directors, professors and students from Department of Public Health or Institutes of Public Health in National Taiwan University, Taipei Medical University, National Taiwan Normal University, National Defense Medical Center, Fu Jen Catholic University, University of Taipei, Tzu Chi University, Chung Shan Medical University, China Medical University, National Cheng Kung University and Kaohsiung Medical University for organizing interviewer team instantly. We thank Dr. Sung-Ching Pan from National Taiwan University Hospital and Dr. Chun-Hsing Liao from Far Eastern Memorial Hospital for executing expert validity of the questionnaires.

## Funding

This project was funding by Taiwan Centers of Disease Control, project LA109025.

## Notes

### Competing Interest Statement

The authors have declared no competing interest.

