## Appendix for "Adherence to preventive behaviors related to Covid-19 in Taiwan in 2020-2021: a population-based follow-up survey"

**Procedures of follow-up surveys**

Participants who agreed to participate in follow-up surveys were contacted during the follow-up surveys. Three waves of follow-up survey were conducted by telephone interview or web-based self-administered questionnaire during the following periods: (1) December 14, 2020 to February 8, 2021, (2) March 27 to April 19, 2021, (3) June 7 to June 14, 2021. Characteristics of participants in the baseline and each follow-up cohort survey is showed as below Appendix Table.

First follow-up

After contacting 2582 participants who agreed to participate follow-up interview in the baseline survey by phone or email, a total of 1629 participants were successfully contacted. Excluding 859 participants who refused to join the follow-up interview or did not complete full questionnaire, 770 participants were enrolled in the 1^st^ follow-up cohort.

Second follow-up

Due to limited resources and interviewers, we contacted those who enrolled in the 1^st^ follow-up cohort and then those refused to participate in 1^st^ follow-up interview with priority to older age. After contacting 2174 participants by phone or email, a total of 1705 participants were successfully contacted. Excluding 849 participants who refused to join the follow-up interview or did not complete full questionnaire, 856 participants were enrolled in the 2^nd^ follow-up cohort.

Third follow-up

The third round only lasted for one week, so we only contacted 1039 participants successfully. Excluding 311 participants who refused to join the follow-up interview or did not complete full questionnaire, 728 participants were enrolled in the 3^rd^ follow-up cohort.

Appendix Table 1.

|  |  | **Baseline** | | **Cohort study** | | | | | | | | | | |
| --- | --- | --- | --- | --- | --- | --- | --- | --- | --- | --- | --- | --- | --- | --- |
|  |  |  |  | 1st follow-up | | | | | 2nd follow-up | | | | 3rd follow-up | |
|  |  | N | % | N | | % | | N | | % | | N | | % |
| **Gender** |  |  |  |  |  | |  | | |  |  |  | | |
|  | Male | 2126 | 44.4 | 328 | 42.65 | | 366 | | | 42.76 | | 298 | | 40.93 |
|  | Female | 2661 | 55.58 | 441 | 57.35 | | 490 | | | 57.24 | | 430 | | 59.07 |
|  | Other | 1 | 0.02 | - | - | | - | | | - | | - | | - |
| **Age group** | |  |  |  |  | |  | | |  | |  | |  |
|  | 20-34 | 1189 | 24.83 | 247 | 32.12 | | 307 | | | 35.86 | | 304 | | 41.76 |
|  | 35-49 | 1366 | 28.53 | 244 | 31.73 | | 246 | | | 28.74 | | 229 | | 31.46 |
|  | 50-64 | 1317 | 27.51 | 190 | 24.71 | | 199 | | | 23.25 | | 89 | | 12.23 |
|  | 65+ | 916 | 19.13 | 88 | 11.44 | | 104 | | | 12.15 | | 106 | | 14.56 |
| **Highest education level** | |  |  |  |  | |  | | |  | |  | |  |
|  | Below elementary school | 386 | 8.06 | 21 | 2.73 | | 25 | | | 2.92 | | 19 | | 2.61 |
|  | Junior high school | 406 | 8.48 | 33 | 4.29 | | 32 | | | 3.74 | | 22 | | 3.02 |
|  | Senior high school | 1172 | 24.48 | 148 | 19.25 | | 141 | | | 16.47 | | 86 | | 11.81 |
|  | College | 2310 | 48.25 | 436 | 56.7 | | 504 | | | 58.88 | | 450 | | 61.81 |
|  | Graduate school | 509 | 10.63 | 131 | 17.04 | | 154 | | | 17.99 | | 151 | | 20.74 |

| Appendix Table 2. Homogeneity test on gender of the expected sample size and baseline cohort data in cities | | | | | | | | |
| --- | --- | --- | --- | --- | --- | --- | --- | --- |
| **City** | **Sample size** | | **Baseline** | | | | ***p*-value** | |
|  | Male | Female | Male | | Female | Total | |  |
| Keelung City | 50 | 50 | 35 | | 50 | 85 | | 0.29 |
| Taipei City | 261 | 297 | 263 | | 276 | 539 | | 0.54 |
| New Taipei City | 416 | 446 | 303 | | 448 | 751 | | <0.01 |
| Taoyuan City | 227 | 236 | 123 | | 210 | 333 | | <0.01 |
| Hsinchu City | 49 | 51 | 50 | | 51 | 101 | | 1 |
| Hsinchu County | 58 | 56 | 58 | | 60 | 118 | | 0.9 |
| Miaoli City | 60 | 56 | 43 | | 47 | 90 | | 0.67 |
| Taichung City | 283 | 301 | 236 | | 300 | 536 | | 0.15 |
| Changhua City | 135 | 133 | 124 | | 137 | 261 | | 0.57 |
| Nantou County | 55 | 52 | 58 | | 57 | 115 | | 0.99 |
| Yunlin County | 75 | 71 | 72 | | 74 | 146 | | 0.82 |
| Chiayi City | 48 | 52 | 49 | | 55 | 104 | | 1 |
| Chiayi County | 58 | 53 | 64 | | 58 | 122 | | 1 |
| Tainan City | 199 | 204 | 154 | | 256 | 410 | | <0.01 |
| Kaohsiung City | 291 | 305 | 256 | | 288 | 544 | | 0.59 |
| Pingtung County | 90 | 88 | 96 | | 107 | 203 | | 0.59 |
| Yilan County | 50 | 50 | 53 | | 51 | 104 | | 1 |
| Hualien County | 50 | 50 | 33 | | 46 | 79 | | 0.35 |
| Taitung County | 51 | 49 | 31 | | 49 | 80 | | 0.14 |
| Penghu County | 51 | 49 | 25 | | 41 | 66 | | 0.13 |
| Note. Sample size was calculated using the population residential data of Department of Household Registration in May 2020 | | | | | | | | |

| Appendix Table 3. Homogeneity test on age group of the expected sample size and baseline cohort data in cities | | | | | | | | | | |
| --- | --- | --- | --- | --- | --- | --- | --- | --- | --- | --- |
| **City** | **Sample size** | | | | **Baseline** | | | | | ***p*-value** |
|  | 20-34 | 35-49 | 50-64 | 65+ | 20-34 | 35-49 | 50-64 | 65+ | Total |  |
| Keelung City | 24 | 27 | 30 | 19 | 19 | 17 | 30 | 19 | 85 | 0.648 |
| Taipei City | 117 | 164 | 151 | 126 | 125 | 166 | 131 | 117 | 539 | 0.637 |
| New Taipei City | 207 | 259 | 243 | 153 | 198 | 223 | 195 | 136 | 752 | 0.667 |
| Taoyuan City | 124 | 146 | 121 | 72 | 73 | 97 | 96 | 67 | 333 | 0.168 |
| Hsinchu City | 24 | 34 | 25 | 17 | 25 | 34 | 25 | 17 | 101 | 1 |
| Hsinchu County | 29 | 38 | 29 | 18 | 29 | 40 | 32 | 17 | 118 | 0.984 |
| Miaoli City | 29 | 32 | 31 | 24 | 18 | 25 | 25 | 22 | 90 | 0.826 |
| Taichung City | 154 | 178 | 156 | 96 | 144 | 159 | 136 | 97 | 536 | 0.867 |
| Changhua City | 69 | 76 | 70 | 53 | 46 | 73 | 83 | 59 | 261 | 0.112 |
| Nantou County | 26 | 27 | 30 | 24 | 25 | 31 | 35 | 24 | 115 | 0.942 |
| Yunlin County | 34 | 40 | 39 | 33 | 35 | 37 | 37 | 37 | 146 | 0.938 |
| Chiayi City | 25 | 28 | 27 | 20 | 26 | 29 | 28 | 21 | 104 | 1 |
| Chiayi County | 25 | 28 | 31 | 27 | 27 | 34 | 33 | 28 | 122 | 0.974 |
| Tainan City | 95 | 117 | 113 | 78 | 127 | 98 | 118 | 67 | 410 | 0.067 |
| Kaohsiung City | 140 | 176 | 164 | 116 | 139 | 161 | 154 | 90 | 544 | 0.594 |
| Pingtung County | 43 | 48 | 51 | 36 | 47 | 51 | 63 | 42 | 203 | 0.95 |
| Yilan County | 25 | 27 | 28 | 20 | 25 | 31 | 29 | 19 | 104 | 0.971 |
| Hualien County | 24 | 27 | 28 | 21 | 20 | 18 | 25 | 16 | 79 | 0.907 |
| Taitung County | 24 | 27 | 29 | 20 | 17 | 19 | 24 | 20 | 80 | 0.838 |
| Penghu County | 27 | 27 | 26 | 20 | 24 | 23 | 18 | 1 | 66 | 0.005 |
| Note. Sample size was calculated using the population residential data of Department of Household Registration in May 2020 | | | | | | | | | | |

Appendix Table 4. Adoption of major preventive behaviors within population who completed all follow-up surveys.

|  |  | Baseline | | Follow-up 1 | | Follow-up 2 | | Follow-up 3 | |  | Friedman test | Wilcoxon Signed Rank test | | |
| --- | --- | --- | --- | --- | --- | --- | --- | --- | --- | --- | --- | --- | --- | --- |
| Items |  | N | % | N | % | N | % | N | % |  | p value |  |  |  |
| Wash hands with water and soap | | |  |  |  |  |  |  |  |  |  |  |  |  |
|  | Never | 5 | 1.61 | 4 | 1.29 | 6 | 1.94 | 1 | 0.32 |  |  |  |  |  |
|  | Seldom | 18 | 5.81 | 17 | 5.5 | 17 | 5.48 | 7 | 2.26 |  |  |  |  |  |
|  | Sometimes | 45 | 14.52 | 60 | 19.42 | 60 | 19.35 | 30 | 9.68 |  |  |  |  |  |
|  | Usually | 98 | 31.61 | 120 | 38.83 | 124 | 40 | 104 | 33.55 |  |  |  |  |  |
|  | Always | 144 | 46.45 | 108 | 34.95 | 103 | 33.23 | 168 | 54.19 |  | **<.0001** | **0.002** | **0.0005** | **<.0001** |
| Wash hands with alcohol sanitizer$ | | |  |  |  |  |  |  |  |  |  |  |  |  |
|  | Never | 27 | 10 | 13 | 4.19 | 13 | 4.19 | 4 | 1.29 |  |  |  |  |  |
|  | Seldom | 30 | 11.11 | 36 | 11.61 | 36 | 11.61 | 15 | 4.84 |  |  |  |  |  |
|  | Sometimes | 49 | 18.15 | 57 | 18.39 | 59 | 19.03 | 28 | 9.03 |  |  |  |  |  |
|  | Usually | 66 | 24.44 | 105 | 33.87 | 105 | 33.87 | 76 | 24.52 |  |  |  |  |  |
|  | Always | 98 | 36.3 | 99 | 31.94 | 97 | 31.29 | 187 | 60.32 |  | **<.0001** | **<.0001** | **<.0001** | **<.0001** |
| Avoid touching eyes and nose | | |  |  |  |  |  |  |  |  |  |  |  |  |
|  | Never | 17 | 5.48 | 11 | 3.55 | 9 | 2.9 | 3 | 0.97 |  |  |  |  |  |
|  | Seldom | 35 | 11.29 | 24 | 7.74 | 22 | 7.1 | 9 | 2.9 |  |  |  |  |  |
|  | Sometimes | 63 | 20.32 | 54 | 17.42 | 68 | 21.94 | 29 | 9.35 |  |  |  |  |  |
|  | Usually | 86 | 27.74 | 118 | 38.06 | 109 | 35.16 | 105 | 33.87 |  |  |  |  |  |
|  | Always | 109 | 35.16 | 103 | 33.23 | 102 | 32.9 | 164 | 52.9 |  | **<.0001** | 0.0744 | 0.0978 | **<.0001** |
| Cover mouth when sneezing and coughing | | | |  |  |  |  |  |  |  |  |  |  |  |
|  | Nor applicable | 111 | 35.81 | 26 | 8.39 | 20 | 6.45 | 30 | 9.68 |  |  |  |  |  |
|  | Never | 5 | 1.61 | 1 | 0.32 | 4 | 1.29 | . | . |  |  |  |  |  |
|  | Seldom | 11 | 3.55 | 3 | 0.97 | 3 | 0.97 | 1 | 0.32 |  |  |  |  |  |
|  | Sometimes | 16 | 5.16 | 17 | 5.48 | 10 | 3.23 | 8 | 2.58 |  |  |  |  |  |
|  | Usually | 36 | 11.61 | 67 | 21.61 | 63 | 20.32 | 59 | 19.03 |  |  |  |  |  |
|  | Always | 131 | 42.26 | 196 | 63.23 | 210 | 67.74 | 212 | 68.39 |  | . | . | . | . |
| Keep indoor social distancing | | |  |  |  |  |  |  |  |  |  |  |  |  |
|  | Never | 22 | 7.1 | 7 | 2.26 | 4 | 1.29 | 3 | 0.97 |  |  |  |  |  |
|  | Seldom | 68 | 21.94 | 33 | 10.65 | 46 | 14.89 | 5 | 1.61 |  |  |  |  |  |
|  | Sometimes | 83 | 26.77 | 100 | 32.26 | 105 | 33.98 | 40 | 12.9 |  |  |  |  |  |
|  | Usually | 91 | 29.35 | 112 | 36.13 | 91 | 29.45 | 131 | 42.26 |  |  |  |  |  |
|  | Always | 46 | 14.84 | 58 | 18.71 | 63 | 20.39 | 131 | 42.26 |  | **<.0001** | **<.0001** | **<.0001** | **<.0001** |
| Keep outdoor social distancing | | |  |  |  |  |  |  |  |  |  |  |  |  |
|  | Never | 14 | 4.52 | 5 | 1.61 | 6 | 1.94 | 2 | 0.65 |  |  |  |  |  |
|  | Seldom | 34 | 10.97 | 23 | 7.42 | 34 | 10.97 | 2 | 0.65 |  |  |  |  |  |
|  | Sometimes | 70 | 22.58 | 76 | 24.52 | 79 | 25.48 | 17 | 5.48 |  |  |  |  |  |
|  | Usually | 115 | 37.1 | 119 | 38.39 | 108 | 34.84 | 110 | 35.48 |  |  |  |  |  |
|  | Always | 77 | 24.84 | 87 | 28.06 | 83 | 26.77 | 179 | 57.74 |  | **<.0001** | **0.0116** | 0.3972 | **<.0001** |
| Wear mask when being unable to keep social distancing | | | | |  |  |  |  |  |  |  |  |  |  |
|  | Never | 6 | 1.94 | . | . | 1 | 0.32 | . | . |  |  |  |  |  |
|  | Seldom | 8 | 2.58 | 3 | 0.97 | 2 | 0.65 | . | . |  |  |  |  |  |
|  | Sometimes | 33 | 10.65 | 9 | 2.9 | 14 | 4.52 | 1 | 0.32 |  |  |  |  |  |
|  | Usually | 84 | 27.1 | 60 | 19.35 | 56 | 18.06 | 8 | 2.58 |  |  |  |  |  |
|  | Always | 179 | 57.74 | 238 | 76.77 | 237 | 76.45 | 301 | 97.1 |  | **<.0001** | **<.0001** | **<.0001** | **<.0001** |
| Rest at home when feeling sick | | |  |  |  |  |  |  |  |  |  |  |  |  |
|  | Nor applicable | 254 | 81.94 | 254 | 81.94 | 241 | 77.74 | 266 | 85.81 |  |  |  |  |  |
|  | Never | 12 | 3.87 | 7 | 2.26 | 15 | 4.84 | 8 | 2.58 |  |  |  |  |  |
|  | Seldom | 5 | 1.61 | 6 | 1.94 | 7 | 2.26 | 3 | 0.97 |  |  |  |  |  |
|  | Sometimes | 5 | 1.61 | 7 | 2.26 | 12 | 3.87 | 3 | 0.97 |  |  |  |  |  |
|  | Usually | 6 | 1.94 | 8 | 2.58 | 5 | 1.61 | 3 | 0.97 |  |  |  |  |  |
|  | Always | 28 | 9.03 | 28 | 9.03 | 30 | 9.68 | 27 | 8.71 |  | . | . | . | . |
| $Delete one sample answered "not applicable" in baseline | | | | |  |  |  |  |  |  |  |  |  |  |

**Cross-sectional survey**

Cross sectional online survey was delivered through the official LINE account of Taiwan CDC. Limited by the default setting of LINE survey, we selected 7 major questions of prevention behaviors from the baseline questionnaire. Two rounds of the online surveys were conducted on Dec 22, 2020 and Jan 28, 2021.

Appendix Table 5.

|  |  | **Baseline** | | **Online cross-sectional survey*** | | | |
| --- | --- | --- | --- | --- | --- | --- | --- |
|  |  |  |  | 1st survey | | 2nd survey | |
|  |  | N | % | N | % | N | % |
| **Gender** |  |  |  |  |  |  |  |
|  | Male | 2126 | 44.4 | 10265 | 21.08 | 13111 | 20.51 |
|  | Female | 2661 | 55.58 | 38340 | 78.72 | 50681 | 79.27 |
|  | Other | 1 | 0.02 | 101 | 0.21 | 140 | 0.22 |
| **Age group** | |  |  |  |  |  |  |
|  | 20-34 | 1189 | 24.83 | 16121 | 33.1 | 22453 | 35.12 |
|  | 35-49 | 1366 | 28.53 | 19110 | 39.24 | 25685 | 40.18 |
|  | 50-64 | 1317 | 27.51 | 10523 | 21.61 | 12590 | 19.69 |
|  | 65+ | 916 | 19.13 | 2952 | 6.06 | 3204 | 5.01 |
| **Residence city** | |  |  |  |  |  |  |
|  | Keelung City | 85 | 1.78 | 677 | 1.39 | 834 | 1.3 |
|  | Taipei City | 539 | 11.26 | 9327 | 19.15 | 11016 | 17.23 |
|  | New Taipei City | 752 | 15.71 | 10682 | 21.93 | 14136 | 22.11 |
|  | Taoyuan City | 333 | 6.95 | 4501 | 9.24 | 8989 | 14.06 |
|  | Hsinchu City | 101 | 2.11 | 1333 | 2.74 | 1801 | 2.82 |
|  | Hsinchu County | 118 | 2.46 | 1103 | 2.26 | 1757 | 2.75 |
|  | Miaoli City | 90 | 1.88 | 631 | 1.3 | 880 | 1.38 |
|  | Taichung City | 536 | 11.19 | 6087 | 12.5 | 7116 | 11.13 |
|  | Changhua City | 261 | 5.45 | 1438 | 2.95 | 1808 | 2.83 |
|  | Nantou County | 115 | 2.4 | 547 | 1.12 | 691 | 1.08 |
|  | Yunlin County | 146 | 3.05 | 641 | 1.32 | 790 | 1.24 |
|  | Chiayi City | 104 | 2.17 | 535 | 1.1 | 648 | 1.01 |
|  | Chiayi County | 122 | 2.55 | 512 | 1.05 | 644 | 1.01 |
|  | Tainan City | 410 | 8.56 | 3231 | 6.63 | 3938 | 6.16 |
|  | Kaohsiung City | 544 | 11.36 | 5084 | 10.44 | 5852 | 9.15 |
|  | Pingtung County | 203 | 4.24 | 894 | 1.84 | 1109 | 1.73 |
|  | Yilan County | 104 | 2.17 | 602 | 1.24 | 822 | 1.29 |
|  | Hualien County | 79 | 1.65 | 475 | 0.98 | 609 | 0.95 |
|  | Taitung County | 80 | 1.67 | 242 | 0.5 | 273 | 0.43 |
|  | Penghu County | 66 | 1.38 | 77 | 0.16 | 102 | 0.16 |
|  | Kinmen County | - |  | 74 | 0.15 | 100 | 0.16 |
|  | Lienchiang County | - |  | 13 | 0.03 | 17 | 0.03 |

Appendix Figure. Trend of preventive behaviors adopted more than often from baseline survey and two cross-sectional survey delivered through LINE.

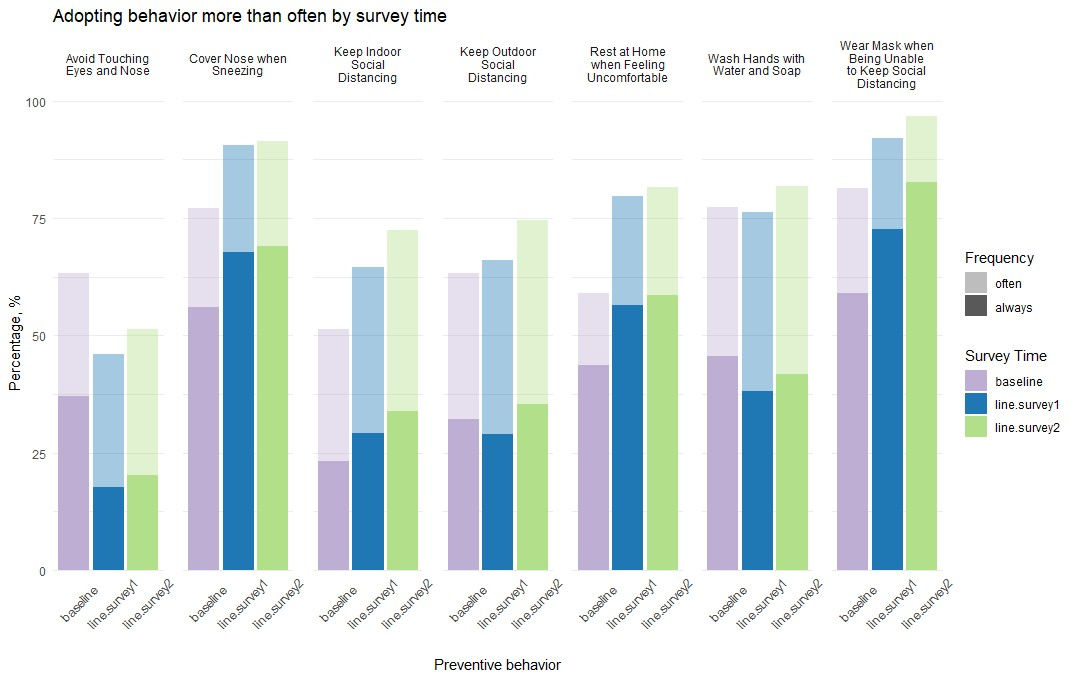

* The execution periods for the following surveys were as follows. Baseline: July 18,2020 to September 16,2020; line.survey1: December 22, 2020 to December 23, 2020; line.survey2: January 28, 2021
